# BURNOUT: A PREDICTOR OF ORAL HEALTH IMPACT PROFILE AMONG NIGERIAN EARLY CAREER DOCTORS

**DOI:** 10.1101/2023.01.16.23284593

**Authors:** OO Ogunsuji, OM Adebayo, KK Kanmodi, OF Fagbule, MA Adeniyi, NT James, AI Yahya, MO Salihu, TO Babarinde, OI Olaopa, TT Selowo, UU Enebeli, DG Ishaya

**Affiliations:** Department of Periodontology & Community Dentistry, University College Hospital, Ibadan, Nigeria; Department of Medicine, University College Hospital, Ibadan, Nigeria; Medical Research Unit, Adonai Hospital, Karu, Nigeria; Department of Community Medicine, Federal Teaching Hospital, Ido-Ekiti, Ekiti State, Nigeria; Department of Obstetrics and Gynaecology, Federal Teaching Hospital, Gombe, Gombe State; Department of Ear, Nose and Throat (ENT), Head and Neck Surgery, University of Maiduguri Teaching Hospital, Maiduguri, Borno State; Department of Behavioural Sciences, University of Ilorin Teaching Hospital Ilorin, Kwara State, Nigeria; Department of Oral and Maxillofacial Surgery, University College Hospital, Ibadan, Nigeria; Department of Chemical Pathology, Jos University Teaching Hospital, Lamingo, Jos, Plateau State, Nigeria; Department of Community Medicine, University of Port Harcourt Teaching Hospital, Port Harcourt, Rivers State, Nigeria; Department of Internal Medicine, Abubakar Tafawa Balewa University Teaching Hospital, Bauchi, Bauchi State, Nigeria

## Abstract

**Introduction:** There have been reported association of oral health disorders with burnout, stress, and mental health. Arguably, with these reported associations, and the current prevalence of burnout amongst Nigerian doctors, exploring the role of burnout on oral health amongst Nigerian doctors is timely. This study aims to determine the relationship between burnout and oral health-related quality of life amongst Early Career Doctors (ECDs) in Nigeria, while also identifying the role other possible predictors plays in this relationship.

**Methods:** This was a cross-sectional study conducted amongst Nigerian ECDs as part of Challenges of Residency Training in Nigeria (CHARTING) II project. A total of 632 ECDs were recruited across thirty-one tertiary hospitals in the 6 geopolitical zones of the country using a multistage cluster sampling technique. A self-administered paper-based semi-structured questionnaire was given to each participant that consented. The tools used to assess burnout and Oral health-related quality of life (OHRQoL) were Copenhagen Burnout Inventory (CBI) and Oral Health Impact Profile (OHIP-14) respectively. Independent samples T-test, ANOVA and Multiple linear regression were used to draw inferences from the data collected.

**Results:** Overall mean OHIP-14 score of all participants was 11.12 (±9.23). The scores for the 3 dimensions of burnout were below 50% with CBI-Personal Burnout having the highest score of 49.96 (±19.15). Significant positive correlations (p < 0.001) were found between OHIP-14 and all the dimensions of burnout, as the burnout scores were increasing, there was a corresponding increase in the OHIP scores thus poorer OHRQoL. The regression model shows that the predictors of OHIP were CBI-PB (p = 0.003), use of fluoride paste (p = 0.039), use of tobacco (p = 0.005) and being a denture user (p = 0.047).

**Conclusion:** This study shows a positive correlation between burnout and OHIP of ECDs. We found that as burnout was increasing, OHIP increased thus implying poorer oral health related quality of life amongst ECDs. The use of fluoride toothpaste, tobacco and denture are other factors we found that could affect the OHIP of ECDs.

## INTRODUCTION

Early in the 21^st^ century, oral health-related disorders were reported as the fourth most expensive condition to manage in most developed countries.^1^ The burden of oral health has been further highlighted by growing evidence of associations between oral health and perennial non-communicable systemic conditions such as cardiovascular diseases, diabetes mellitus.^2,3^ Furthermore, increase in prevalence of oral health disorders such as bruxism, oral lichen planus, temporomandibular joint disorders, aphthous ulcers, dry mouth, periodontal diseases and caries have been positively associated with stress and mental health.^4-6^

A systematic review of burnout among Nigerian doctors reported burnout ranged from 23.6% to 51.7%.^7^ Studies carried out among resident doctors in Nigeria reported high level of stress and exhaustion due to excessive working hours, time pressure, having to deal with patients. Other reasons were preparation and thoughts of exams and demands of family life.^8, 9^ Burnout and stress appears to have been used interchangeably in most of the reported literature amongst Nigerian doctors. Stress, though not well defined compared to burnout with clearly defined dimensions, is often highly correlated to burnout especially in emotional exhaustion dimension.^10-13^

Oral-health-related quality of life (OHRQoL) has been defined as ‘the impact of oral disease and disorders on aspects of everyday life that a patient or person values, that are of sufficient magnitude, in terms of frequency, severity or duration to affect their experience and perception of their life overall’.^14, 15^ The concept of OHRQoL appeared only in the early 1980s in contrast to the general health-related quality of life (HRQOL) which had been popular about two decades earlier. The perception of the impact of oral diseases on systemic health was perhaps responsible for this delay with some researchers then often rejecting the idea that oral diseases could be related to general health.^16^

Oral health-related quality of life is used increasingly as a person-reported outcome measure in oral health research. This use requires detailed understanding of the factors that influence and those which may act to confound or mediate the relationship between clinical status and OHRQoL. OHRQoL is an important health outcome, and an understanding of its determinants is necessary to inform the design and evaluation of interventions to enhance it. OHRQoL is a multidimensional construct that reflects amongst other things, people’s comfort when sleeping, eating, and engaging in social interaction, their self-esteem, and their satisfaction with respect to their oral health. OHRQoL is associated with functional factors, psychological factors, social factors and experience of pain or discomfort.

Though evidence suggests a relationship between burnout and oral disorders, there is dearth of information on the relationship between burnout and oral health. Furthermore, the relationship between burnout and oral health along with oral health related quality of life among Nigerian doctors especially the early career doctors has not been previously reported. Similarly, the role of confounders such as sociodemographic factors, dental care utilization and oral maintenance behavior has been sparingly accounted for in existing literature on stress and oral health. This study aims to determine the relationship and correlation between burnout and oral health-related quality of life, while also identifying the role other possible predictors plays in this relationship.

## METHODOLOGY

### Study design, location, and participants

This was a cross-sectional study conducted amongst Nigerian Early Career Doctors (ECDs) as part of Challenges of Residency Training in Nigeria (CHARTING) II project. The protocol for this project has already been described in a previous publication.^17^ The study was conducted in a total of 31 tertiary hospitals in Nigeria. Respondents included in this study were ECDs who had been employed for at least 3 months in their hospital. Non-ECDs such as principal medical and dental officers (PMO/PDO) and consultants were excluded from the study. All ECDs provided written informed consent to be recruited into the study. Ethical approval was obtained from the National Health Research Ethics Committee of Nigeria (NHREC/01/01/2007-06/08/2020B).

### Sample size and sampling technique

Sample size was calculated based on the assumption of 50% prevalence of burnout amongst ECDs. Non-response rate of 10% and design effect was equally factored in based on the clusters. A sample size of 679 was arrived at using StatCalc^®^ of EpiInfo7 produced by the Centre for Disease Control and Prevention. During data cleaning, participants with missing variables were removed and a total of 632 respondents were left.

A multistage cluster sampling technique was used, and 31 centers were randomly selected using simple random sampling technique (balloting) from a list of all centres across the 6 geopolitical zones as follows: Northwest (5), Northcentral (7), Northeast (3), Southsouth (5), Southwest (9) and Southeast (2). Five to ten departments were then randomly selected using a simple random sampling technique (balloting) from a list of all the departments in each Centre. All willing and qualified participants in the selected departments were recruited.

### Instrument and data collection

Prior to the conduct of the study, the questionnaire which was composed of standardized and validated study instruments was pretested amongst 57 ECDs (who were not part of the study participants) to ensure the questions were simple, concise, unambiguous and reflects all the options available to the respondents. Participants were given the questionnaire to fill twice within a 48-hour interval. The intra-respondent intra-class correlation coefficient for the Oral Health Impact Profile Score was 0.927 (92.7%), subsequently the questionnaire was adapted and adopted for use. The questionnaire was a self-administered paper-based semi-structured which was divided into 4 sections. Section one covered the sociodemographic characteristics of participants. Section 2 was on the oral health practices and other factors which were based on the WHO Oral Health Survey methods.^18^ Section 3 was on Copenhagen Burnout Inventory tool which had already been validated amongst early career doctors in Nigeria.^19^ The final section was on the Oral Health Impact Profile (OHIP-14), a tool for assessing the oral health-related quality of life, which is a shortened form of the original OHIP-49.^20^

#### Oral hygiene practices

To categorize the extent of sugar consumption by each participant, the response to each of the seven questions to determine sugar consumption was assigned scores ranging from 0 for “never” to 5 for “several times a day” with a maximum score of 35 obtainable. Scores within “0 – 10”, “11 – 20” and “21 – 35” were categorized as low, moderate, and high consumption respectively.^21^ Alcohol consumption was dichotomized based on if an individual had taken alcohol within the last 30 days or not. Similarly, tobacco use was dichotomized based on status of either smoking/chewing tobacco in any form or not.

#### Copenhagen Burnout Inventory

CBI comprised of 19 positively and negatively framed items that cover three dimensions: Personal burnout (6 items), Work-related burnout (7 items) and Patient-related burnout (6 items). Each question has a 5-point Likert scale (never, seldom, sometimes, often, always) response option which study participants picked from. Before calculating the mean scores for each of the components, the negatively worded question was reversed. Weighted scores (percentages) were assigned to each option as follows; never (0), seldom (25), sometimes (50), often (75) and always (100).^22^ The score for the set of questions on each of the three dimensions was added and the average of the scores was calculated. The higher the percentage score within a dimension, the more susceptible an individual is to burnout within that dimension.

#### Oral Health Impact Profile (OHIP-14)

A shortened form of OHIP-49 which consists of 14 questions measuring the oral health-related quality of life in 7 domains (each domain has 2 questions). Possible responses to each question are assigned scores ranging from 0 for ‘never’ to 4 for ‘very often’. These scores are then summed up, the maximum score for each domain is 8, and the maximum overall score obtainable is 56, indicating worse OHRQoL as the OHIP score increases.^23^

### Data Management and analysis

The independent variables include sociodemographic variables, oral health practices and the Copenhagen Burnout Inventory (CBI) dimensions namely personal burnout (CBI-PB), work-related burnout (CBI-WRBO) and patient-related burnout (CBI-PRBO) and OHIP-14 was the dependent variable.

Data was analyzed with IBM SPSS version 26. Continuous variables such as OHIP-14, CBI and Age were found to be normally distributed using Shapiro Wilk’s test (p > 0.05), as such their mean and standard deviation were reported. The frequency and percentages of all categorical variables were reported. Bivariate analyses to test for the difference in the OHIP-14 scores across each categorical variable were carried out using either Independent samples t-test or One way ANOVA. Scores for the 3 dimensions of CBI were correlated with OHIP-14 scores using Pearson’s correlation coefficient. Finally, OHIP-14 scores were regressed on the significant independent variables from the bivariate analyses to get a predictive model. Significance level was set at p < 0.05.

## RESULTS

### Sociodemographic and Oral health practices

A total of 632 ECDs were recruited, with a median age of 32 (29 – 36) years. Almost two-thirds of respondents were males (65.5%). A larger proportion of the respondents were married (55.5%) and are registrars (46.0%) **Table 1**. Approximately 5.2% of the respondents use tobacco, with a similar proportion of respondents also using dentures. About 15.3% of ECDs recruited used chewing sticks to clean their teeth, while only 3.5% said they were not using toothbrush. Over one-third of the participants approximately 40% do not practice interdental cleaning (**Table 2**).

**TABLE 1:**
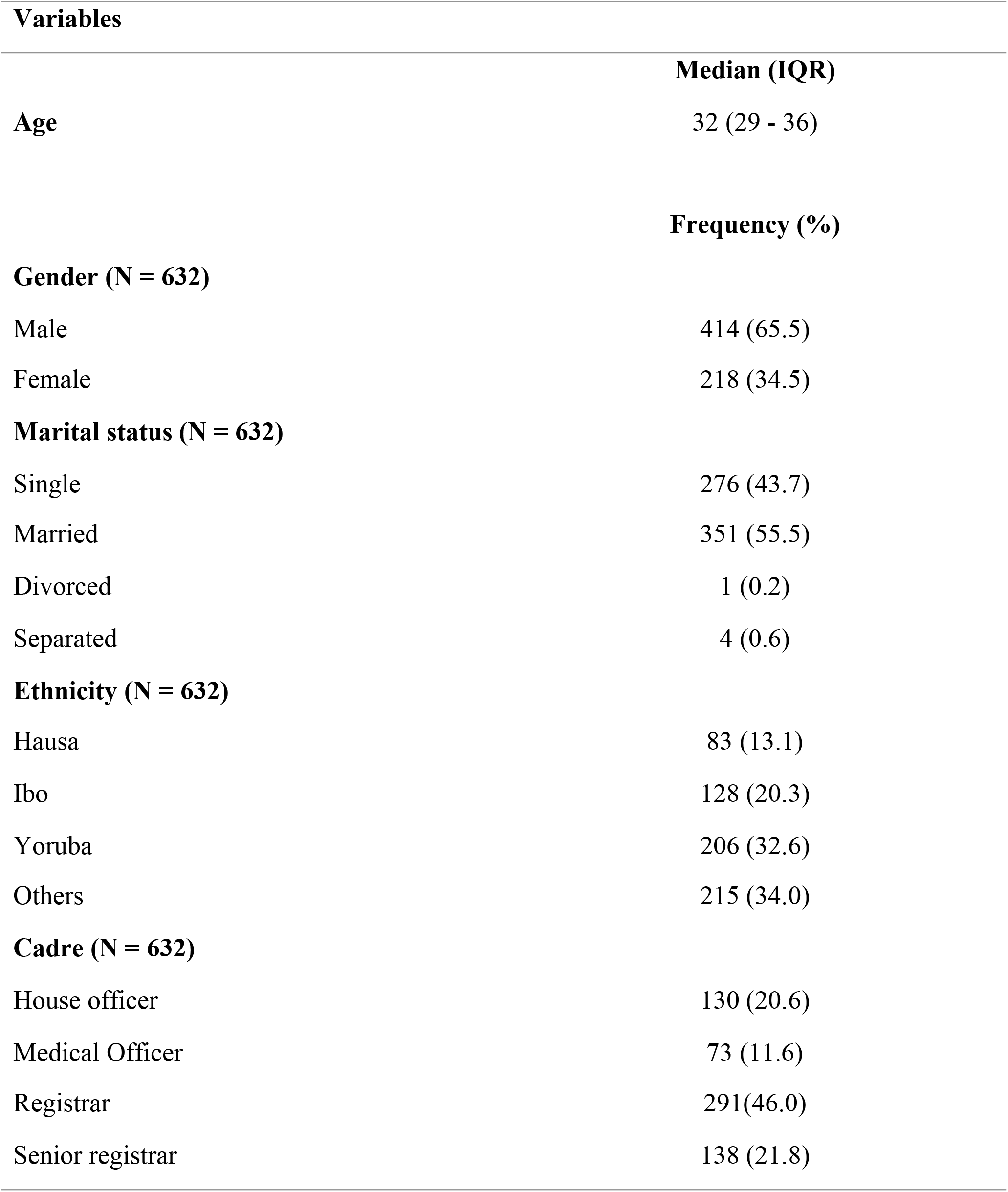
Sociodemographic Characteristics of ECDs (N = 632)

| **Variables** |  |
| --- | --- |
|  | **Median (IQR)** |
| **Age** | 32 (29 - 36) |
|  | **Frequency (%)** |
| **Gender (N = 632)** |  |
| Male | 414 (65.5) |
| Female | 218 (34.5) |
| **Marital status (N = 632)** |  |
| Single | 276 (43.7) |
| Married | 351 (55.5) |
| Divorced | 1 (0.2) |
| Separated | 4 (0.6) |
| **Ethnicity (N = 632)** |  |
| Hausa | 83 (13.1) |
| Ibo | 128 (20.3) |
| Yoruba | 206 (32.6) |
| Others | 215 (34.0) |
| **Cadre (N = 632)** |  |
| House officer | 130 (20.6) |
| Medical Officer | 73 (11.6) |
| Registrar | 291(46.0) |
| Senior registrar | 138 (21.8) |

**TABLE 2:** Oral hygiene and Behavioral practices among ECDs.

| **Variables** | **Frequency (%)** |
| --- | --- |
| **Denture use** |  |
| No | 599 (94.8) |
| Yes | 33 (5.2) |
| **Frequency of tooth cleaning** |  |
| Maximum of once daily | 394 (62.3) |
| 2 or more times daily | 238 (37.7) |
| **Use of chewing stick** |  |
| No | 535 (84.7) |
| Yes | 97 (15.3) |
| **Use of toothbrush** |  |
| No | 22 (3.5) |
| Yes | 610 (96.5) |
| **Interdental cleaning** |  |
| No | 255 (40.3) |
| Yes | 377 (59.7) |
| **Texture of brush** |  |
| Hard | 83 (13.1) |
| Medium | 456 (72.2) |
| Soft | 81 (12.8) |
| Don't know | 12 (1.9) |
| **Use of Fluoride paste** |  |
| No | 22 (3.5) |
| Yes | 551 (87.2) |
| Don't know | 59 (9.3) |
| **Tobacco** |  |
| No | 599 (94.8) |
| Yes | 33 (5.2) |
| **Alcohol consumption (30 days)** |  |
| No | 132 (20.9) |
| Yes | 500 (79.1) |
| **Refined sugar consumption** |  |
| Low | 181 (28.6) |
| Moderate | 390 (61.7) |
| High | 61 (9.7) |
| **Previous dental visit** |  |
| No | 276 (43.7) |
| Yes | 356 (56.3) |

### OHIP and Burnout

**Table 3** showed the overall mean OHIP-14 score of all participants was 11.12 (±9.23). The highest score was obtained in the psychological discomfort domain (2.26) closely followed by the physical disability domain (2.17), while the lowest score was obtained in the functional limitation domain (0.61). Question on “being self-conscious because of state of the teeth or mouth” had the highest score of (1.35), while the questions on “the sense of taste worsening” and “trouble pronouncing words” had the lowest scores of 0.28 and 0.33 respectively.

**Table 3:** OHIP-14 Domain and Overall scores among the respondents.

| **OHIP Domains** | **Mean (SD)** | **Mean (SD)** |
| --- | --- | --- |
| **Functional limitation** |  | 0.61 (1.10) |
| Difficulty pronouncing words | 0.33 (0.70) |  |
| Felt worsened sense of taste | 0.28 (0.61) |  |
| **Physical pain** |  | 1.56 (1.77) |
| Painful aching | 0.83 (1.01) |  |
| Uncomfortable to eat | 0.74 (0.96) |  |
| **Psychological discomfort** |  | 2.26 (2.16) |
| Self-conscious | 1.35 (1.37) |  |
| Felt tense | 0.91 (1.04) |  |
| **Physical disability** |  | 2.17 (2.10) |
| Diet has been unsatisfactory | 1.04 (1.09) |  |
| Interrupted meals | 1.15 (1.21) |  |
| **Psychological disability** |  | 1.91 (1.93) |
| Difficulty to relax | 1.04 (1.12) |  |
| Little embarrassed | 0.88 (1.00) |  |
| **Social disability** |  | 1.72 (1.73) |
| Felt irritable with others | 0.95 (1.01) |  |
| Difficulty doing usual jobs | 0.78 (0.90) |  |
| **Handicap** |  | 1.07 (1.47) |
| Less satisfaction | 0.70 (0.92) |  |
| Unable to function | 0.36 (0.72) |  |
| **Overall OHIP-14 score** |  | 11.12 (9.23) |

The scores for the 3 dimensions of burnout were below 50% with CBI-PB having the highest score of 49.96 (±19.15) and the CBI-PRBO having the lowest score of 26.21 (±20.16) **Figure 1**. Significant positive correlations (p < 0.001) were found between OHIP-14 and all the dimensions of burnout, as the burnout scores were increasing, there was a corresponding increase in the OHIP scores thus poorer OHRQoL. The highest correlation was found between OHIP and CBI-PB (r = 0.273), while the least correlation was found between OHIP and CBI-PRBO (r = 0.163) **Table 4**.

**Figure 1:**
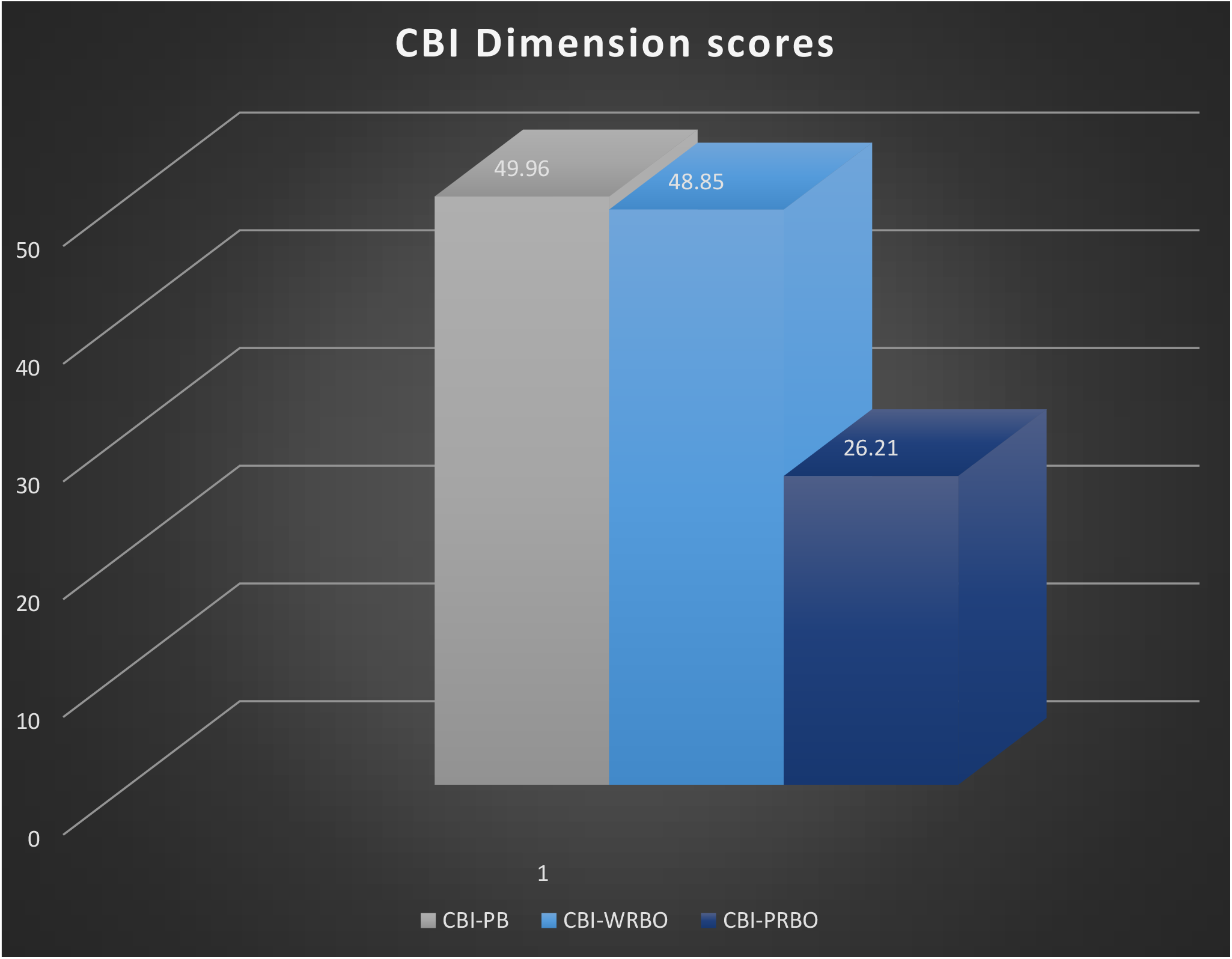
Copenhagen Burnout Inventory Dimensions.

**Table 4:** Correlation between OHIP and Copenhagen burnout dimensions of ECDs.

|  | **OHIP** | **CBI-PB** | **CBI-WRBO** | **CBI-PRBO** |
| --- | --- | --- | --- | --- |
| **OHIP** | 1 |  |  |  |
| **CBI-PB** | 0.273* | 1 |  |  |
| **CBI-WRBO** | 0.244* | 0.782* | 1 |  |
| **CBI-PRBO** | 0.163* | 0.393* | 0.415* | 1 |

*\*Correlation is significant at 0.01 level ( $p < 0.01$ )*

### OHIP association with ECDs sociodemographic and oral hygiene practices

Across the different cadres of ECDs, house officers significantly had the highest OHIP score (worst OHRQoL), while senior residents had the least OHIP score, thus better OHRQoL. ECDs using denture had higher OHIP score thus worse OHRQoL compared to those not using denture. Likewise, ECDs who used toothbrush and fluoride containing toothpaste significantly had lower OHIP score and thus better OHRQoL compared to ECDs who do not use toothbrush or fluoride containing toothpaste. **Table 5**

**Table 5:** Association of OHIP with ECDs sociodemographic and oral hygiene practices.

| **Variables** | **Mean OHIP (± SD)** | **P-value** |
| --- | --- | --- |
| **Age** |  |  |
| ≤35 years | 11.24 (9.58) | 0.587 |
| > 35 years | 10.82 (8.25) |  |
| **Gender** |  |  |
| Male | 11.00 (9.33) | 0.633 |
| Female | 11.37 (9.04) |  |
| **Marital status** |  |  |
| Single | 11.21 (9.85) | 0.859 |
| Married | 11.07 (8.74) |  |
| **Ethnicity** |  |  |
| Hausa | 12.88 (10.08) | 0.136 |
| Ibo | 11.84 (9.05) |  |
| Yoruba | 10.49 (9.30) |  |
| **Cadre** |  |  |
| House Officer | 13.28 (10.86) | 0.016* |
| Medical officer | 11.07 (9.70) |  |
| Registrar | 10.81 (8.43) |  |
| Senior Registrar | 9.79 (8.65) |  |
| **Denture Use** |  |  |
| No | 10.91 (8.99) | 0.049* |
| Yes | 15.10 (12.30) |  |
| **Frequency of cleaning** |  |  |
| Not more than once | 11.00 (8.86) | 0.668 |
| 2 or more times daily | 11.33 (9.82) |  |
| **Chewing stick** |  |  |
| No | 11.26 (9.12) | 0.395 |
| Yes | 10.39 (9.81) |  |
| **Use of toothbrush** |  |  |
| No | 15.59 (9.61) | 0.021* |
| Yes | 10.96 (9.18) |  |
| **Interdental cleaning** |  |  |
| No | 11.38 (9.09) | 0.568 |
| Yes | 10.95 (9.33) |  |
| **Brush texture** |  |  |
| Medium | 10.81 (9.28) | 0.172 |
| Others | 11.93 (9.06) |  |
| **Use of fluoride paste** |  |  |
| Yes | 10.80 (8.98) | 0.005* |
| No | 17.23 (13.02) |  |
| Don't know | 11.90 (9.23) |  |
| **Tobacco** |  |  |
| Yes | 17.54 (12.07) | 0.001* |
| No | 10.77 (8.92) |  |
| **Alcohol use** |  |  |
| Yes | 11.27 (8.99) | 0.836 |
| No | 11.09 (9.30) |  |
| **Sugar consumption** |  |  |
| Low | 9.64 (8.32) | 0.045* |
| Moderate | 11.40 (9.03) |  |
| High | 13.79 (12.04) |  |
| **Previous dental visit** |  |  |
| No | 10.33 (9.14) | 0.043* |
| Yes | 11.74 (9.26) |  |

*\*Statistically significant at 0.05 level ( $p < 0.05$ )*

### Predictors of OHIP

OHIP-14 was regressed on all the significant variables from **table 5** along with the 3 dimensions of CBI. The regression model (**Table 6)** shows that the predictors of OHIP were CBI-PB (p = 0.003), use of fluoride paste (p = 0.039), use of tobacco (p = 0.005) and being a denture user (p = 0.047).

**TABLE 6:** Multiple Regression analysis (dependent variable is OHIP-14 score)

| Variables | Unstandardized<br>beta coefficient | p-value | 95% CI |  |
| --- | --- | --- | --- | --- |
|  |  |  | *Lower* | *Upper* |
| Constant | 6.556 | 0.033 | 0.541 | 12.570 |
| CBI-PB | 0.090 | 0.003 | 0.031 | 0.148 |
| Use fluoride paste (Ref DON'T USE) | -4.398 | 0.039 | -8.584 | -0.212 |
| Use tobacco (Ref DON'T USE) | 5.222 | 0.005 | 1.552 | 8.892 |
| Denture user (Ref DON'T USE) | 3.196 | 0.047 | 0.046 | 6.345 |
| ***F (14, 617) = 6.074, P < 0.001 \*Ref stands for reference group\**** |  |  |  |  |

Equation for the model is presented below

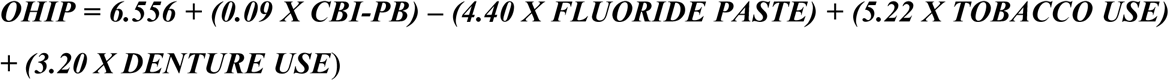

For a unit increase in CBI-PB, an ECDs OHIP score will significantly increase by 0.09. ECDs who use fluoride toothpaste will have their OHIP score reduce significantly by almost 4.5units compared to those not using fluoride paste. ECDs using tobacco will have their OHIP increase significantly by 5.22 units compared to those not using tobacco. Finally, ECDs who are denture users will have their OHIP increase significantly by 3.20 units compared to those not wearing denture.

## DISCUSSION

This study reported the mean OHIP value of our respondents to be 11.12, this finding is similar to the mean OHIP values reported by other Nigerian researchers though their studies were in different population groups.^24-26^ Our mean OHIP score in this study suggests low frequency or degree of severity of oral health problems amongst ECDs.^27^ ECDs reported their highest score in the psychological discomfort and physical disability domains, this finding contrasted with the studies earlier mentioned, where the highest scores reported amongst their respondents was in the physical pain domain. ECDs by nature of their knowledge as health professionals are perhaps better equipped in knowing what to do to prevent painful oral health conditions such as caries and periodontitis thus have fewer episodes of toothache. They may have other forms of oral health conditions such as dental appearance, use of appliances (dentures, orthodontic appliances), lost dentition leading to relatively more psychological discomfort compared to pain.

The trend of CBI dimension scores in this study were similar to those reported in the pilot study conducted to validate CBI or OLBI in comparison to MBI.^19^ The strong positive correlation between CBI-PB and CBI-WRBO was equally similar to the findings from the pilot study.^19^ This study found that only CPI-PB dimension could predict Oral health related quality of life amongst ECDs, the higher the CPI-PB score, the worse the OHRQoL. One would have expected the CBI-WRBO to also be a significant predictor, but this is not the case. This makes a strong case for the CBI-PB dimension as the most important dimension of burnout and relates to how an average ECD values their own physical and mental health and gives it higher priority ahead of patients or work. This study found the use of tobacco and denture as predictors, they both increased OHIP score by over 5 and 3 units respectively when all other factors have been controlled for. The effect of tobacco on oral health has been reported by studies.^28, 29^ Tobacco has reportedly led to qualitative difference in the microbiological constituents of plaque found in current smokers and former smokers compared to never smokers.^30^ Its potential to impair several aspects of innate and immune response tips the homeostatic balance towards exaggerated tissue breakdown and poorer oral health outcomes. Increased collagenolytic activity, suppressed gingival inflammation due to reduction in the number of gingival vessels have reportedly led to impaired and delayed healing in smokers compared to non-smokers following therapy.^30^

Denture wearers in this study had worse OHRQoL, similar to the study by Allen and McMillan.^31^ The presence of a tooth reportedly influences how individuals perceive themselves,^31^ this perhaps explains why the ECDs using dentures had higher OHIP scores and the most affected domain was psychological discomfort.

This study found the use of fluoride toothpaste among ECDs to be a protective factor in lowering the OHIP score. This was similar to the finding by Leon *et al*, where the use of fluorides in management of caries led to a positive impact on OHRQoL.^32^ The role of fluoride within the oral cavity helps in the formation of fluorapatite which reduces the solubility of hydroxyapatite crystals during acid attack and helps prevent caries.^33, 34^ Furthermore, fluoride also inhibits bacterial enzyme activities, thus reducing acid production and preventing the dissolution of the oral tissues, thus leading to improved oral health outcomes.^33, 34^

One of the limitations of this study was our reliance on the self-report by the ECDs. To limit information bias arising from this, the instrument used for collecting information (questionnaire) used was pretested, standardized, validated for ECDs. Most of the questions were precise and close ended, and the Copenhagen burnout tool also used different questions to test the same hypothesis. Furthermore, while this study tried to identify the predictors of OHIP, by virtue of the design, the relationship between OHIP and Burnout (using CBI) was what we determined and not the causation or temporal sequence.

Notwithstanding these limitations, this study had its strengths. First, this study is believed to be the first study to explore the relationships between oral health and burnout among ECDs in Nigeria. Second, this study was a nationally representative study, as it surveyed ECDs across different hospitals in the six geopolitical zones in Nigeria. Third, this study adopted the use of standardized instruments as its data collection tool; hence, the data obtained from this study is reliable.

In conclusion, the findings from this study show there is a relationship between burnout and oral health-related quality of life amongst ECDs. We found that as burnout was increasing, the oral health impact increased with poorer oral health related quality of life amongst ECDs. The use of fluoride toothpaste, tobacco and denture are other factors we found that could affect the OHIP of ECDs.

## Data Availability

The data that support this article are not publicly available. The data contains information that could compromise the privacy of research participants but are available from the corresponding author (Ugo Enebeli) upon reasonable request.

## Conflict of Interest

All authors are members of NARD as at the time of this study; however, the study was independently conducted and reported. NARD played only a funder’s role.

## Funding

This work was supported by the Nigerian Association of Resident Doctors (NARD) under funding for Research & Statistics Committee (Grant No. 0002).

## Author Contributions

OOO and AOM contributed to conceptualization; OOO, AOM, KKK and FOF contributed to methodology; All authors contributed to data collection; OOO contributed to data analysis; AOM contributed to administration; OOO, KKK and FOF contributed to writing of initial draft; all authors contributed to writing, review, and editing of final draft; all the authors listed were involved in the writing of the final draft, and all the authors agreed to be responsible for the outcome of the manuscript.

## Notes

### Competing Interest Statement

The authors have declared no competing interest.

### Funding Statement

The work herein presented was supported by the Nigerian Association of Resident Doctors (NARD) under funding for Research & Statistics Committee (Grant No. 0002), with Website: https://nardrcn.com.ng/. NARD played only a funding role.

### Author Declarations

Ethics committee that approved this study: National Health Research Ethics Committee of Nigeria. Approval number: NHREC/01/01/2007-06/08/2020B. Form of consent obtained: Written informed consent was obtained from all consenting participants.

## REFERENCES

1. Petersen PE, Bourgeois D, Ogawa H, Estupinan-Day S, Ndiaye C. The global burden of oral diseases and risks to oral health. Bulletin of the World Health Organization. 2005;83(9):661–669.

2. Nazir MA. Prevalence of periodontal disease, its association with systemic diseases and prevention. International Journal of Health Sciences. 2017;11(2):72–80.

3. Vasiliou A, Shankardass K, Nisenbaum R, Quiñonez C. Current stress and poor oral health. BMC Oral Health. 2016;16(1):1–8.

4. Kandagal V, Shenai P, Chatra L, Ronad Y, Kumar M. Effect of stress on oral mucosa. Biol Biomed Rep. 2012;1(1):13–16.

5. Finlayson TL, Williams DR, Siefert K, Jackson JS, Nowjack-Raymer R. Oral health disparities and psychosocial correlates of self-rated oral health in the National Survey of American Life. American Journal of Public Health. 2010;100(S1):S246–S255.

6. Armfield JM, Mejía GC, Jamieson LM. Socioeconomic and psychosocial correlates of oral health. International Dental Journal. 2013;63(4):202–209.

7. Ogunsuji O, Adebayo O, Olaopa O, Efuntoye O, Agbogidi J, Kanmodi K, et al. Burnout among Nigerian Doctors: a systematic review. The Nigerian Medical Practitioner. 2019;76(1-3):24–29.

8. Adeolu JO, Yussuf OB, Popoola OA. Prevalence and Correlates of job stress among junior doctors in the University College Hospital, Ibadan. Annals of Ibadan Postgraduate Medicine. 2016;14(2):92–98.

9. Ndom R, Makanjuola A. Perceived stress factors among resident doctors in a Nigerian teaching hospital. West African Journal of Medicine. 2004;23(3):232–235.

10. Rothenberger DA. Physician burnout and well-being: a systematic review and framework for action. Diseases of the Colon & Rectum. 2017;60(6):567–576.

11. Freudenberger HJ. Staff Burnout. Journal of Social Issues. 1974;30(1):159–165.

12. Maslach C, Jackson SE. The measurement of experienced burnout. Journal of Organizational Behavior. 1981;2(2):99–113.

13. Maslach C, Jackson SE, Leiter MP. The Maslach Burnout Inventory Manual second edition. Consulting Psychologists Press. 1986(June 2015).

14. Gururatana O, Baker SR, Robinson PG. Determinants of children’s oral-health-related quality of life over time. Community Dent Oral Epidemiol. 2014;42(3):206–215.

15. Brennan DS, Spencer AJ. Dental visiting history between ages 13 and 30 years and oral health-related impact. Community Dentistry and Oral Epidemiology. 2014;42(3):254–262.

16. Bennadi D, Reddy CVK. Health-Related Quality of Life. Journal of International Society of Preventive and Community Dentistry. 2013;3(1):1–6.

17. Ez UA, Tolani MA, Adeniyi MA, Ogbonna VI, Isokariari O, Igbokwe MC, et al. Challenges of residency training and early career doctors in Nigeria Phase II: Update on objectives, design, and rationale of study. Nigerian Journal of Medicine. 2020;29(4):714–719.

18. WHO Oral Health Surveys - Basic Methods. 5th Edition 2013.p.1–137

19. Ogunsuji O, Ogundipe H, Adebayo O, Oladehin T, Oiwoh S, Obafemi O, et al. Internal Reliability and Validity of Copenhagen Burnout Inventory and Oldenburg Burnout Inventory Compared with Maslach Burnout Inventory among Nigerian Resident Doctors: A Pilot Study. Dubai Medical Journal. 2022;5(2):89–95.

20. Slade GD. Derivation and validation of a short-form oral health impact profile. Community Dentistry and Oral Epidemiology. 1997;25(4):284–290.

21. Ogunsuji O, Dosumu E, Dairo M, Ogunsuji A. Self assessment of oral health and risk factors affecting oral hygiene status in adolescents attending dental clinic in University College Hospital, Ibadan. Annals of Ibadan Postgraduate Medicine. 2021;19(1):70–77.

22. Kristensen TS, Borritz M, Villadsen E, Christensen KB. The Copenhagen Burnout Inventory: A new tool for the assessment of burnout. Work & Stress. 2005;19(3):192–207.

23. Mary AV, Mahendra J, John J, Moses J, Ebenezar AR, Kesavan R. Assessing quality of life using the oral health impact profile (OHIP-14) in subjects with and without orthodontic treatment need in Chennai, Tamil Nadu, India. Journal of Clinical and Diagnostic Research. 2017;11(8):ZC78.

24. Braimoh OB, Alade GO. Oral health-related quality of life and associated factors of elderly population in Port Harcourt, Nigeria. Saudi Journal of Oral Sciences. 2019;6(1):18–22.

25. Lawal FB, Taiwo JO, Arowojolu MO. How valid are the psychometric properties of the oral health impact profile-14 measure in adult dental patients in Ibadan, Nigeria? Ethiopian Journal of Health Sciences. 2014;24(3):235–242.

26. Isiekwe G, Onigbogi O, Olatosi O, Sofola O. Oral health quality of life in a Nigerian university undergraduate population. Journal of the West African College of Surgeons. 2014;4(1):54.

27. Ikebe K, Watkins CA, Ettinger RL, Sajima H, Nokubi T. Application of short-form oral health impact profile on elderly Japanese. Gerodontology. 2004;21(3):167–176.

28. Fernandes MJ, Ruta DA, Ogden GR, Pitts NB, Ogston SA. Assessing oral health-related quality of life in general dental practice in Scotland: validation of the OHIP-14. Community Dentistry and Oral Epidemiology. 2006;34(1):53–62.

29. Sagtani RA, Thapa S, Sagtani A. Smoking, general and oral health related quality of life–a comparative study from Nepal. Health and Quality of Life Outcomes. 2020;18(1):1–7.

30. Lalla E, Papapanou PN. Modifying Factors. In:Lang NP, Lindhe J.(eds.) Clinical Periodontology and Implant Dentistry. 6^th^ edition. West Sussex, UK: John Wiley & Sons Ltd; 2015.p.278–279.

31. Allen PF, McMillan AS. The impact of tooth loss in a denture wearing population: an assessment using the Oral Health Impact Profile. Community Dental Health. 1999;16(3):176–180.

32. León s, Rivera M, Payero S, Correa-Beltrán G, Hugo FN, Giacaman RA. Assessment of oral health-related quality of life as a function of non-invasive treatment with high-fluoride toothpastes for root caries lesions in community-dwelling elderly. International Dental Journal. 2019;69(1):58–66.

33. Kanduti D, Sterbenk P, Artnik B. Fluoride: a review of use and effects on health. Materia Socio-Médica. 2016;28(2):133.

34. O Mullane D, Baez R, Jones S, Lennon M, Petersen P, Rugg-Gunn A, et al. Fluoride and oral health. Community Dental Health. 2016;33(2):69–99.

